# Mobile robotic systems in patient-facing functions: national acceptability survey, single site feasibility study and cost-effectiveness analysis

**DOI:** 10.1101/2020.09.30.20204669

**Authors:** Peter R Chai, Farah Z Dadabhoy, Hen-wei Huang, Jacqueline N Chu, Annie Feng, Hien M Le, Joy Collins, Marco da Silva, Marc Raibert, Chin Hur, Edward W Boyer, Giovanni Traverso

## Abstract

**Objective:** To understand the acceptability of patient-facing mobile robotic systems on a national scale, conduct a pilot feasibility study to deploy and measure satisfaction associated with clinical evaluation using a mobile telehealth robot in the emergency department (ED) and to build a decision analytic model to gauge the potential of a robotic system to prevent COVID-19 infections and conserve personal protective equipment in the ED.

**Design:** Mixed study comprising an online sampling-based survey, single-site observational clinical trial and development of a decision analytic model.

**Setting:** A quaternary care, urban, academic, emergency department in Boston, Massachusetts, USA.

**Participants:** For the acceptability survey, we recruited N=1000 individuals living in the United States participating in an online sampling from the survey provider YouGov. In the ED study, we enrolled 40 individuals over 18 years old presenting to the ED for evaluation.

**Interventions:** In the pilot ED study, consenting participants were exposed to a mobile robotic system facilitated triage interview controlled by an emergency medicine clinician. Afterwards, participants completed a survey to measure their satisfaction with the robotic system.

**Main outcome measures:** Acceptability of mobile robot facilitated tasks in healthcare (national survey), satisfaction with interaction of a robotic system (ED study), number of potential SARS-CoV-2 infections avoided and cost savings (US dollars) per year per ED (decision analytic model).

**Results:** In the national survey, participants rated the use of robotics for a variety of patient-facing healthcare functions useful or very useful. The perceived usefulness increased when asked to consider these functions in the context of the COVID-19 pandemic. In the ED, 40 patients completed study procedures; 92.5% (N=37) reported satisfaction with the robotic system. Most participants (82.5%, N=33) reported their experience being evaluated by a robotic system was as good as an in-person encounter. Our decision analytic model estimated that robotic evaluations could prevent 2.68 infections per ED yearly and save $1 million annually per ED by decreasing PPE and additional staffing in a triage space.

**Conclusions:** Robotic systems were broadly acceptable across the US and their acceptance increased in the setting of COVID-19. Mobile robotic-enabled teleheath facilitated contactless evaluation of ED patients and was highly acceptable and equivalent to an in-person history. Robotic platforms may prevent healthcare-associated COVID-19 transmission to healthcare workers and have a significant cost savings if widely implemented among healthcare systems.

**Trial Registration:** Clinicaltrials.gov, NCT04452695

**Summary Box:** *What is already known on the topic:*

- Mobile robots can provide telemedicine services that enable remote evaluation of patients in various settings.
- The use of mobile systems may facilitate contactless evaluation of patients while minimizing use of personal protective equipment (PPE) during the COVID-19 pandemic.

*What this study adds:*

- Our study suggests that there is broad acceptability of mobile robotic systems to perform patient-facing tasks in the hospital. This sentiment is reflected in satisfaction of patients in the emergency department interacting with a mobile robotic teletriage system during our open label pilot study.
- Our decision analysis model suggests that adoption of robotic systems for teletriage in the emergency department may prevent up to 2.68 SARS-CoV-2 infections among healthcare workers over one year per emergency department in the United States as well as a cost savings in deploying a robotic system compared to a clinician to evaluate patients during the COVID-19 pandemic.

## Introduction

The COVID-19 pandemic has changed the manner in which healthcare workers (HCW) interact with patients. Personal protective equipment (PPE), social distancing and special triage facilities to screen symptomatic individuals have been implemented to protect HCWs and prevent transmission of SARS-CoV-2.^1–6^ Despite these measures, HCWs continue to be a population with high risk for COVID-19 disease; in Italy, up to 20% of infections were HCW.^7^ The consequences of SARS-CoV-2 infection among HCW extend beyond the morbidity and mortality from sequelae of COVID-19. HCW who acquire COVID-19 disease are unable to provide direct patient care, thereby decreasing the availability of a vital workforce during the pandemic.^8,9^

While advances towards development of pharmacotherapy and vaccines to address COVID-19 disease continue to advance at a rapid rate, many healthcare systems have settled into new procedures to maximize social distancing to minimize the potential for SARS-CoV-2 transmission. In order to accomplish this, many hospital systems have expanded telehealth capabilities to limit contact with patients who may have COVID-19 disease.^3^ These telehealth solutions enable providers to deliver care virtually, determine the need for additional testing and potentially conduct follow up visits in a contactless manner.^10,11^ A key advantage of telehealth systems is both the decreased utilization of PPE and a decreased risk of HCW and potentially patient transmission of SARS-CoV-2. On March 20, 2020, the United States Food and Drug Administration (FDA) issued an emergency use authorization (EUA) that allows for the adoption of technologies to facilitate patient monitoring and telemedicine without the traditional lengthy approval process.^12^

While many of these telehealth platforms rely upon static, patient controlled tablet computers or smartphones, a robotic telehealth system controlled by a clinician facilitates a dynamic evaluation process that can be deployed in the hospital setting.^13^ Mobile telemedicine systems can facilitate evaluation of patients in various settings.^14^ For example, robotic systems may permit a mobile telepresence that can move between high-risk patients, rooms or wards in a hospital setting.^15^ In field hospitals erected to manage the influx of patients with COVID-19, the use of an agile robotic system may obviate the need to install temporary static infrastructure that is needed to support traditional telemedicine systems.^16^ With continued focus on decreasing exposure to HCW and reducing PPE utilization, a robotic system could help facilitate triage and evaluation of individuals with COVID-19 in the hospital setting.^17^ Prior to widespread implementation of robotic systems to provide patient care during the COVID-19 pandemic, it is important to understand the acceptability of these systems by patients as well as the potential economic impact to a healthcare system that adopts robotic systems for teletriage.^18^ In this investigation, we sought to understand attitudes towards robotic-facilitated healthcare tasks like telemedicine, contactless vital signs and nasal and oral swabs through a national sampling of individuals in the United States. Additionally, we deployed a robotic system to facilitate contactless assessment of patients with potential COVID-19 in the emergency department.

Finally, we developed a decision analytic model to estimate the potential decrease in SARS-CoV-2 transmission with a robotic system to facilitate triage in an emergency department in the setting of the COVID-19 pandemic.

## Methods

Our investigation consisted of three parts: 1. A survey of N=1000 individuals across the U.S. to understand attitudes towards robotic systems in medical care; 2. an open label trial of a mobile robotic telehealth system to facilitate triage and telemedicine in the emergency department during the COVID-19 pandemic in Boston, Massachusetts; 3. a decision analytic model to understand the overall cost savings of deploying a robotic system within emergency departments across the United States. Mass General Brigham Institutional Review (IRB) approval was received prior to initiating the study (MGB 2020P000957).

### Patient and public involvement

Patients and the public were involved in the open label trial portion of the research through screening as they arrived at the emergency department at Brigham and Women’s Hospital. Research questions around patient satisfaction were developed by the study investigators using the Teleheatlh Usability Questionnaire as a template.^19^ Patients and the public were not involved in the design of the study although they were informed that results of their participation would be submitted in the form of a peer-reviewed manuscript.

### National survey on attitudes towards robotics systems

We partnered with YouGov (London, UK), a global market research and data analytics service to conduct a national survey of United States residents on attitudes towards robotic systems in healthcare. We developed a survey questionnaire modeled off the Negative Attitudes Towards Robots Scale (NARS), a platform agnostic quantitative measure that evaluates attitudes towards interactive robotic system.^20,21^ Survey responses were measured using a 5-Point Likert scale from strongly disagree to strongly agree. We also developed questions surrounding the use of robotic systems to facilitate healthcare-related tasks like telemedicine, contactless vital sign assessment, performing a nasal/oral swab, IV catheter placement, phlebotomy and turning a patient in bed (See supplementary information). We asked these questions in the context of interaction with robotic systems in the hospital. Next, we asked participants to consider the use of robotic systems in the context of the COVID-19 pandemic, and whether they would be accepting of robotics to mitigate the use of PPE. Survey responses were measured using a 5-Point Likert scale from useless to extremely useful.

Participants were surveyed from August 18, 2020 to August 21, 2020. YouGov utilizes a sample matching technique that selects potential participants from its internal repository of potential survey respondents matching characteristics of the United States Census. Next, matching is verified using a Mahalanobis distance metric to ensure the recruited sample from the YouGov database corresponds with general population characteristics. Raw results were tabulated, and weights were applied to ensure representation of a national sample. We measured a composite NARS score among study participants using the Subordinate “S1” (Negative Attitude toward Situations of Interactions with Robots) Scale. This scale describes baseline negative attitudes towards robotic systems. We calculated basic descriptive statistics (mean, standard deviation, and minimum and maximum values) to characterize NARS scores amongst participants. For questions considering usefulness of robotic systems in performing hospital tasks, we calculated basic descriptive statistics to compare the average scores of usefulness (median and interquartile range) considering the contexts of a regular healthcare setting and COVID-19. Next, we used the Wilcoxon Signed Rank test to compare the average response rates between these two contexts to see if the differences were statistically significant. Data analysis was completed using STATA/IC 16.1.

### Patient acceptance of a robotic system in the emergency department

We conducted an open label pilot descriptive trial to understand the feasibility and acceptability of a robotic platform to conduct telehealth triage within the emergency department during the COVID-19 pandemic (NCT04452695). Our study was conducted in the emergency department of a large, urban, academic tertiary care center. The Brigham and Women’s Hospital Emergency Department evaluates approximately 60,000 patients annually. During the COVID-19 pandemic, standard waiting room triage systems were altered to accommodate individuals with upper respiratory symptoms consistent with potential COVID-19 infection. A large 25 feet by 45 feet tent was erected outside the ED waiting room to evaluate patients who present with potential COVID-19 infection. Patients enter this space, receive initial triage by a nurse, and then a rapid screening history by a clinician. This screening helps determine whether individuals are tested for SARS-CoV-2 by institutional and state guidelines. Otherwise well-appearing patients are then tested and discharged home.

We deployed Spot, an agile, quadruped robotic platform (Boston Dynamics, Waltham MA) to perform contactless clinician history in individuals presenting to the ED during the COVID-19 pandemic (Figure 1).^22^ This system consisted of a four-legged robot outfitted with a secure radio communication relay to a tablet controller that allows a single operator to navigate the robot. The robot carries stereo cameras that permit obstacle avoidance and payload cameras that allow the robot to be remotely navigated. We also outfitted the robot with a tablet computer running a real-time person-to-person video link that allowed us to conduct telemedicine encounters in the emergency department. We initially used a bluetooth-linked commercial speaker to augment the audio from the tablet computer however, we discovered that due to an inconsistent connection, the use of the native tablet speakers was adequate for patient communication. Providers were outfitted with a set of noise cancelling headphones to facilitate patient interviews. We conducted a standardized training program to instruct emergency medicine providers (physician assistants and physicians) to operate the robotic system and tablet computer.

**Figure 1:**
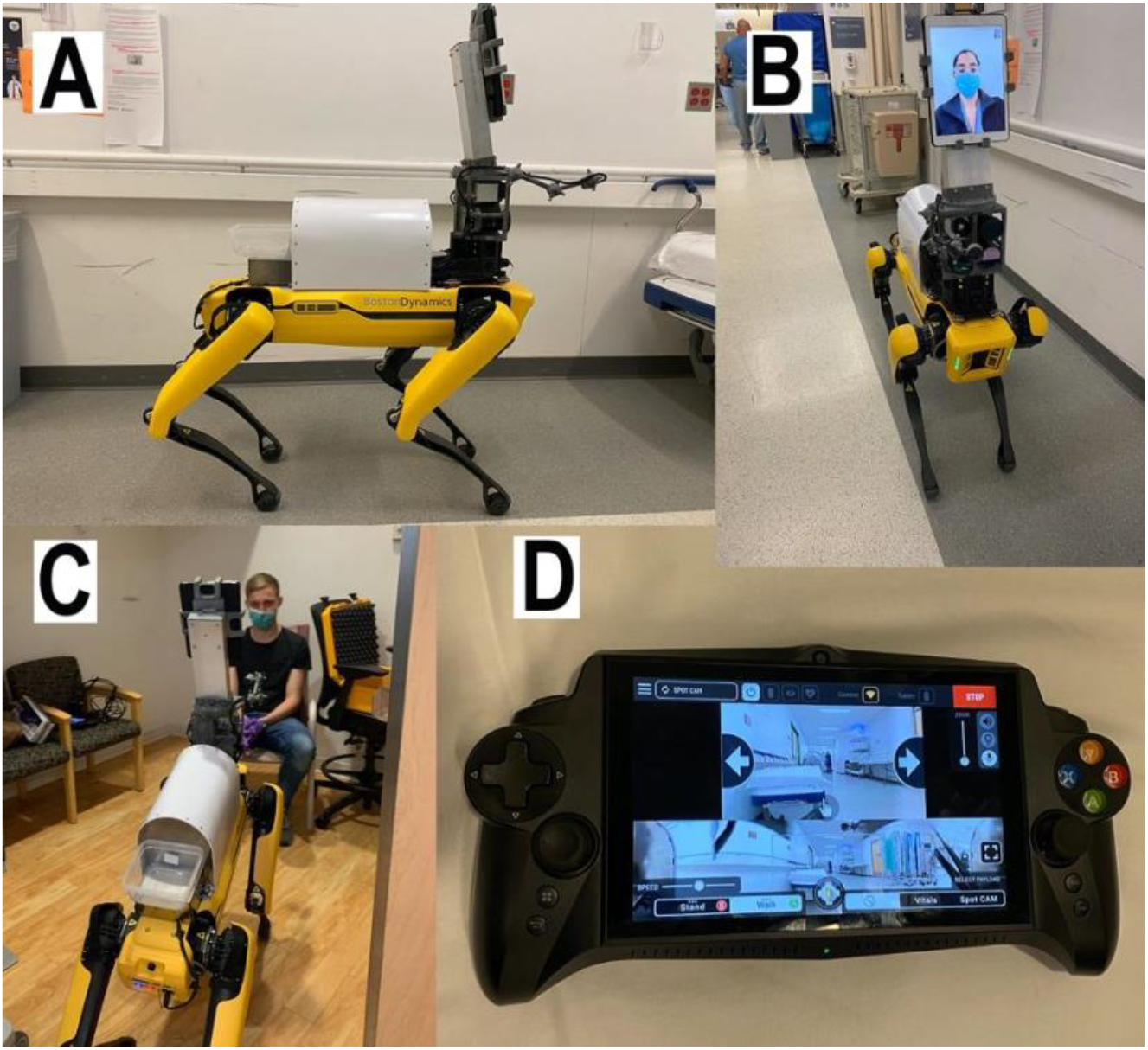
Images of Dr. Spot (Boston Dynamics, Waltham MA) demonstrating mobile robotic system (A), outfitted with tablet computer to facilitate face-to-face evaluation by emergency department providers (B/C). Dr. Spot is controlled using a custom Android-based interface and controller (D).

We enrolled patients presenting to the emergency department, triaged either in the novel tent space, the standard ED waiting room, or directly roomed into the ED between April 15, 2020 and August 5, 2020. Participants verbally consented to participate in the study. Next, participants were exposed to the robotic teletriage system controlled by a trained emergency medicine provider. The provider both navigated the robot through the emergency department to the participant’s triage space or ED room, and obtained a history via the integrated video link on the tablet computer. Questions and counseling were completed at the discretion of the provider. At the conclusion of the encounter, participants completed a quantitative assessment regarding their experience with the robotic system. The robotic system chassis was sterilized with ethanol wipes in between each encounter. The quantitative assessment was developed using the Telehealth Usability Questionnaire as a guide.^19^ We utilized RedCAP, a secure, web-based survey instrument, to collect survey data.^23^ Spot was decontaminated using standard alcohol-based hospital wipes in between each participant encounter.

Participant data were downloaded from RedCAP. We calculated basic descriptive statistics to describe participant attitudes and the user response to the robotic teletriage system. All data analysis was completed in STATA v15.

### Decision analytic model of robotic systems in the emergency department

To estimate the potential impact of the robotic teletriage system on infections and cost, we developed a decision analytic model using TreeAge (Williamstown, MA). We generated a decision tree to compare the strategies of robotic teletriage versus standard HCW triage (Supplementary Figure 1). We estimated the probabilities of patients presenting to the ED being infected with SARS-CoV-2 and transmitting SARS-CoV-2 to HCW (if infected) or becoming infected with SARS-CoV-2 from HCW (if not infected) over an 8-hour ED shift and scaled the results over 1,000 shifts (approximately one year) (Supplementary Table 2). Given significant uncertainty regarding rate of COVID transmission in the healthcare setting, we performed sensitivity analyses on SARS-CoV-2 transmission probabilities based on best estimates from the literature. We calculated the number of potential healthcare-acquired SARS-CoV-2 transmissions via standard triage to estimate the upper bound of preventable cases by robotic triage. Cost of standard triage included an extra emergency medicine physician’s salary over an 8 hour shift (given that the robotic strategy is estimated to expedite triage twice as fast) and additional PPE required. The robotic strategy included a wide range of possible estimates for robot cost based on currently available technology ($500 to $100,000).

#### Role of the funding source

The funding sources were not involved in study design, data collection, analysis or drafting of the manuscript. Boston Dynamics generously donated Dr. Spot, the robot used in this investigation and technical support during the study period.

## Results

### National survey on acceptance of robotic systems in healthcare

We enrolled N=1000 individuals participating in the YouGov survey repository. Individuals represented a national sampling. Mean age of participants was 48.7; 46.5 % (N=465) were male and 53.5% (N=535) were female. Thirty-five percent reported finishing college or graduate school (Table 1). The mean NARS S1 score for survey participants was 16.3 with a standard deviation of 4.8. Scores range from 6-30, with those scoring in the lower range closer to 6 having a less negative attitude towards situations involving interactions with robots than those scoring in the higher range closer to 30.^21^Twenty-five percent of participants scored at or below a score of 13 and seventy-five percent at or below a score of 19. A mean score of 16.3 falls within the lower range of NARS S1 indicating that the study population was relatively accepting of interactions with robots.

**Table 1:**
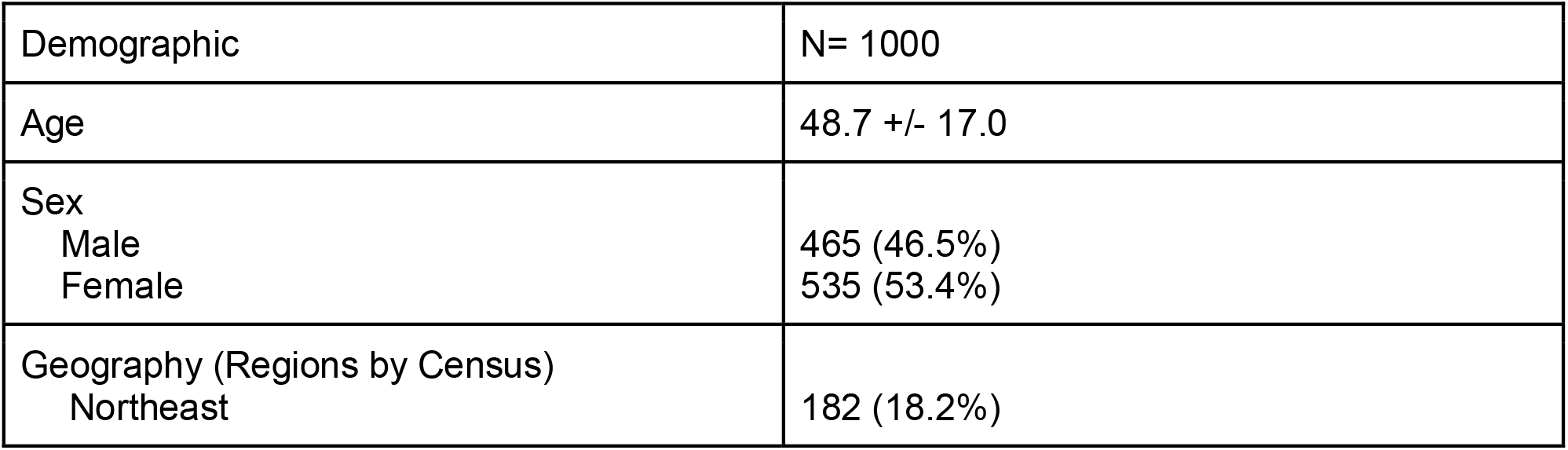
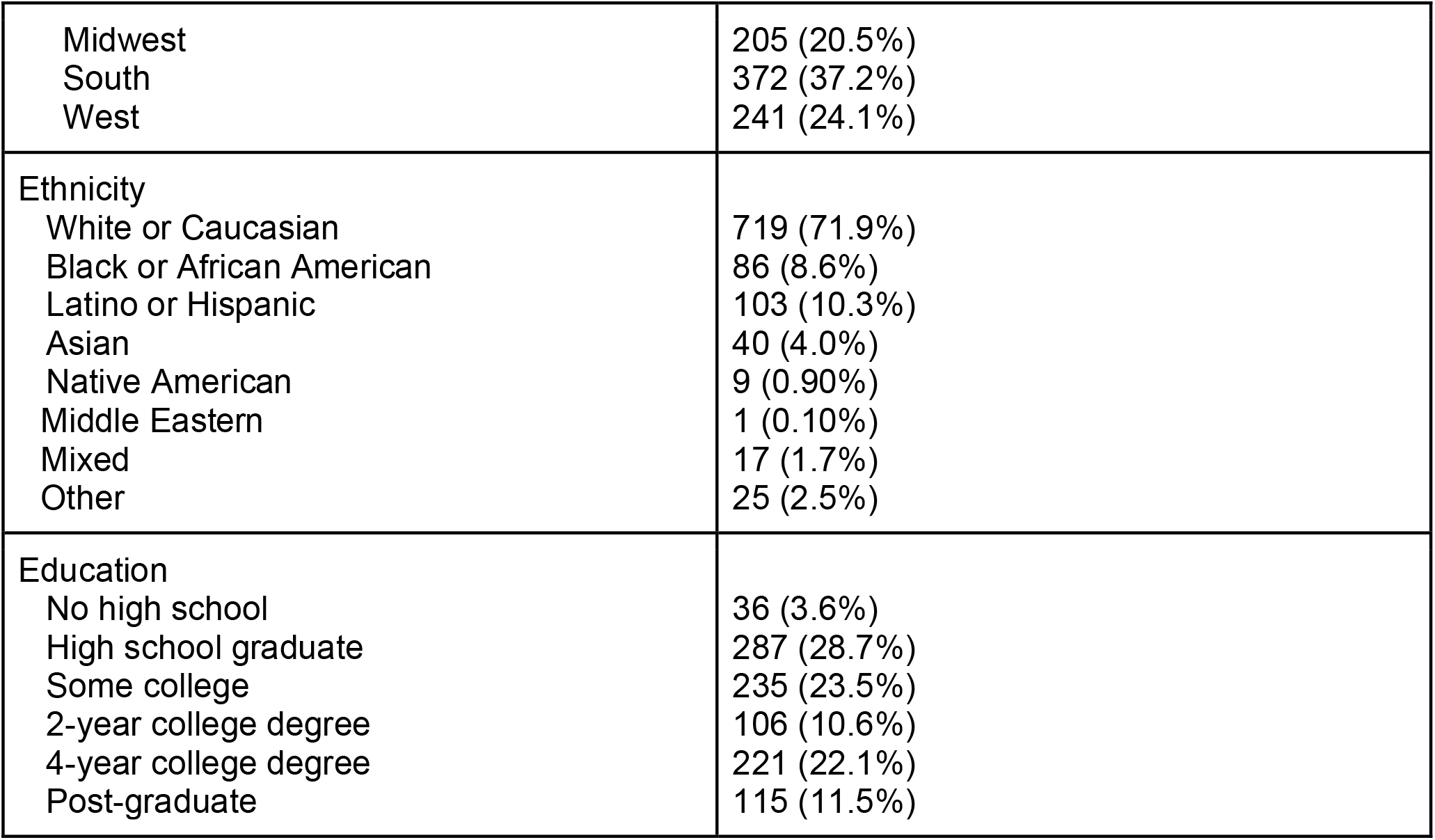
Demographics from National US Survey

We selected six questions that reflected care tasks that robotic systems may assist during the COVID-19 pandemic: Telemedicine evaluation, acquiring contactless vital signs, placing an intravenous catheter, performing phlebotomy, nasal and oral swabs, and turning a patient in bed (proning an individual) (Table 2). Median scores for these tasks performed in a hospital ranged from a 3 (placing an IV, phlebotomy, nasal and oral swabs) to 4 (telemedicine, contactless vitals, turning a patient in bed). When asked to consider the use of robotic systems to perform these same tasks in the hospital during the current COVID-19 pandemic, median score for performing nasal and oral swabs changed from 3 (neutral) to 4 (somewhat useful). Other median scores were unchanged.

**Table 2:** Participant response around usefulness of robotic systems for tasks in the hospital

|  | Usefulness of Robotic Systems for hospital tasks | Usefulness of Robotic Systems for hospital tasks in context of COVID-19 | p-value |
| --- | --- | --- | --- |
| Facilitating telemedicine interview with physician or nurse |  |  | 0.1574 |
| 1-Extremely Useless | 68 (6.8%) | 77 (7.7%) |  |
| 2-Somewhat Useless | 94 (9.4%) | 90 (9.0%) |  |
| 3-Not Useful/Useless | 178 (17.8%) | 168 (16.8%), |  |
| 4- Somewhat Useful | 373 (37.3%) | 351 (35.1%) |  |
| 5-Extremely Useful | 287 (28.7%) | 314 (31.4%) |  |
| *Median Score* | 4 | 4 |  |
| Acquisition of contactless vital signs |  |  | 0.0003 |
| 1-Extremely Useless | 46 (4.6%) | 76 (7.6%) |  |
| 2-Somewhat Useless | 62 (6.2%) | 62 (6.2%) |  |
| 3-Not Useful/Useless | 129 (12.9%) | 123 (12.3%) |  |
| 4- Somewhat Useful | 350 (35.0%) | 336 (33.6%) |  |
| 5-Extremely Useful | 413 (41.3%) | 403 (40.3%) |  |
| *Median Score* | 4 | 4 |  |
| Obtaining nasal or oral swabs<br>1-Extremely Useless<br>2-Somewhat Useless<br>3-Not Useful/Useless<br>4- Somewhat Useful<br>5-Extremely Useful<br>*Median Score* | 120 (12.0%)<br>168 (16.8%)<br>213 (21.3%)<br>307 (30.7%)<br>192 (19.2%)<br>3 | 116 (11.6%)<br>163 (16.3%)<br>195 (19.5%)<br>287 (28.7%)<br>239 (23.9%)<br>4 | 0.0023 |
| Placing an intravenous catheter<br>1-Extremely Useless<br>2-Somewhat Useless<br>3-Not Useful/Useless<br>4- Somewhat Useful<br>5-Extremely Useful<br>*Median Score* | 190 (19.0%)<br>210 (21.0%)<br>213 (21.3%)<br>228 (22.8%)<br>159 (15.9%)<br>3 | 167 (16.7%)<br>198 (19.8%)<br>199 (19.9%)<br>228 (22.8%)<br>208 (20.8%)<br>3 | <0.001 |
| Performing phlebotomy<br>1-Extremely Useless<br>2-Somewhat Useless<br>3-Not Useful/Useless<br>4- Somewhat Useful<br>5-Extremely Useful<br>*Median Score* | 170 (17.0%)<br>197 (19.7%)<br>217 (21.7%)<br>249 (24.9%)<br>167 (16.7%)<br>3 | 152 (15.2%)<br>194 (19.4%)<br>203 (20.3%)<br>236 (23.6%)<br>215 (21.5%)<br>3 | <0.001 |
| Helping position or turn a patient in bed<br>1-Extremely Useless<br>2-Somewhat Useless<br>3-Not Useful/Useless<br>4- Somewhat Useful<br>5-Extremely Useful<br>*Median score* | 59 (5.9%)<br>63 (6.3%)<br>136 (13.6%)<br>371 (37.1%)<br>371 (37.1%)<br>4 | 64 (6.4%)<br>91 (9.1%)<br>136 (13.6%)<br>331 (33.1%)<br>378 (37.8%)<br>4 | 0.0425 |
| N=1000<br>*Survey scores using a 5-Point Likert Scale |  |  |  |

While median scores did not change for most tasks, the Wilcoxon Sign Rank Test demonstrated that a statistically significant number of individuals increased their usefulness ranking of robotic systems in the context of the COVID-19 pandemic. For the tasks of acquiring contactless vital signs and turning a patient in bed the average differences were statistically significant (p-values= 0.0003, 0.0425), with more patients considering them useful in the general hospital setting. For the tasks of conducting a nasal/oral swab, placing an intravenous catheter, and performing phlebotomy the average differences were statistically significant (p-values= 0.0023, <0.001, <0.001), with more patients considering them useful for the context of COVID-19. There was no significant change in the usefulness of robotic systems for telemedicine. This suggests that for certain hospital tasks participants who felt that robotic systems could be useful felt they could be even more useful in the context of COVID-19 (Table 2).

### Emergency department satisfaction after robotic teletriage

Over the study period, N=51 potential participants were approached. Forty-one consented and we enrolled N=40 participants (Figure 2). Forty participants who enrolled in the study successfully completed the quantitative assessment. One participant was unable to be enrolled due to technical difficulties associated with the operation of Spot. Mean age of participants was 45.8; 27.5% (N=11) were male, 72.5% (N=29) were female. 45% reported completing college or graduate school (Table 3).

**Table 3:** Study demographics

| Demographic | N= 40 |
| --- | --- |
| Age | 45.8 +/- 2.7 |
| Sex<br>Male<br>Female | 11 (27.5%)<br>29 (72.5%) |
| Ethnicity<br>Caucasian<br>African American<br>Latino or Hispanic<br>Asian<br>Other | 22 (55%)<br>7 (17.5%)<br>9 (22.5%)<br>2 (5%)<br>1 (2.5%) |
| Education<br>Less than high school<br>High school<br>Some college<br>College degree<br>Some graduate school<br>Trade school<br>Graduate degree | 5 (12.5%)<br>5 (12.5%)<br>12 (30%)<br>11 (27.5%)<br>2 (5%)<br>2 (5%)<br>3 (7.5%) |

**Figure 2:**
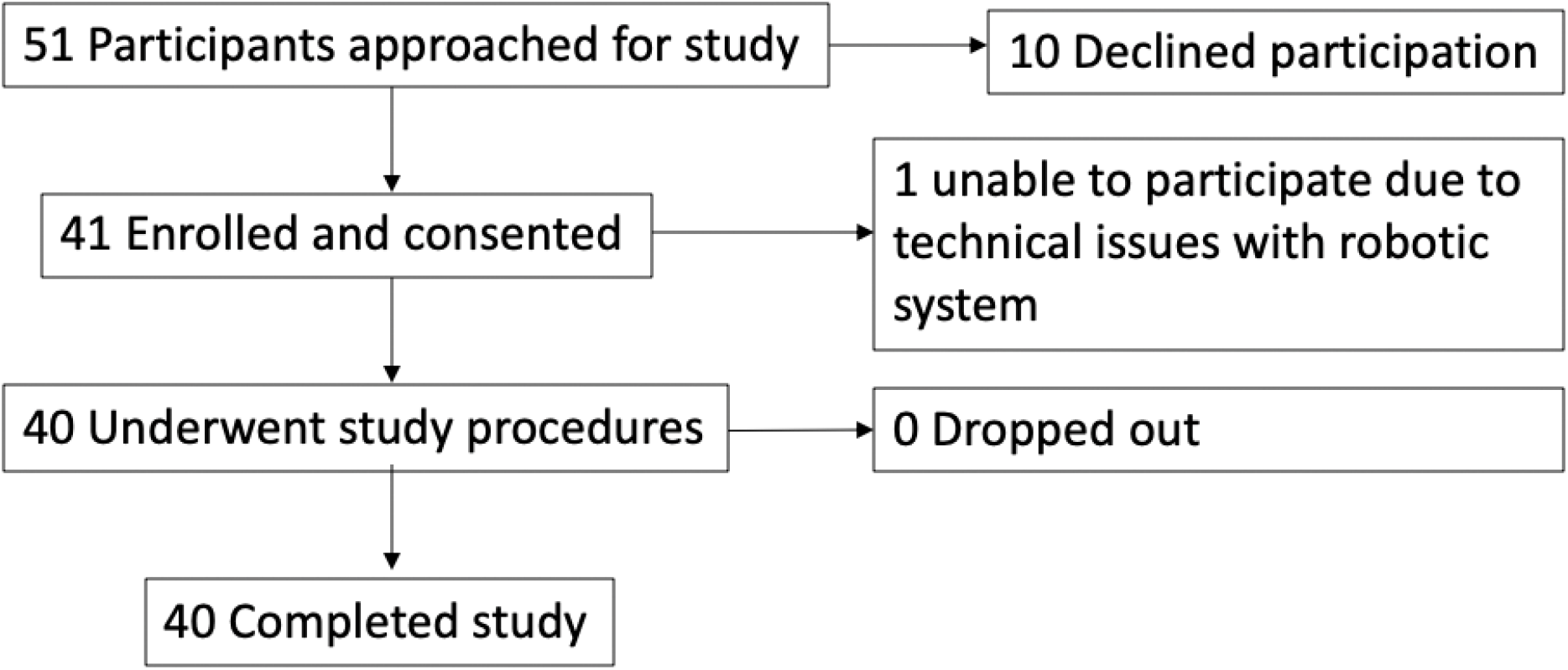
Study enrollment schema.

Most participants reported being satisfied with the robotic system (N=37, 92.5%), and were also satisfied with their interaction with clinicians facilitated by this system (N=34, 85%) (Table 4). In parallel, participants considered the quality of video provided by the robotic system to be adequate (N=38, 95%). Despite experiencing an ED environment that can be loud and chaotic, especially during the COVID-19 pandemic, the on-board audio quality was adequate for participants to understand questions and interact with clinicians (N=35, 87.5%). Interacting with a robotic system, although novel for participants, was intuitive, and felt simple.

Importantly, participants considered their robot-facilitated interaction with a clinician to be as good as an in-person encounter (N=33, 82.5%). They reported that their clinician was able to provide adequate information that was understandable despite not being present in the triage space with a provider (N=35, 87.5%). When asked about future healthcare-related visits, participants considered virtual care facilitated by a robotic platform acceptable (N=34, 85%), and reported they would be willing to interact with a robotic system in the future (N=37, 92.5%).

**Table 4:** User response to the robotic telemedicine system

| Variable | N= 40 |
| --- | --- |
| Overall satisfaction with robotic system<br>Dissatisfied<br>Neutral<br>Satisfied | 0<br>3 (7.5%)<br>37 (92.5%) |
| Interaction with clinician using robotic system<br>Dissatisfied<br>Neutral<br>Satisfied | 0 (0%)<br>6 (15%)<br>34 (85%) |
| Video quality of robotic system<br>Dissatisfied<br>Neutral<br>Satisfied | 0 (0%)<br>2 (5%)<br>38 (95%) |
| Audio quality of robotic system<br>Dissatisfied<br>Neutral<br>Satisfied | 2 (5%)<br>3 (7.5%)<br>35 (87.5%) |
| Interaction as good as in-person encounter<br>Disagree<br>Neutral<br>Agree | 5 (12.5%)<br>2 (5%)<br>33 (82.5%) |
| Information provided by clinician using robotic platform<br>Dissatisfied<br>Neutral<br>Satisfied | 0 (0%)<br>3 (7.5%)<br>37 (92.5%) |
| Comfort interacting with a clinician using a robotic platform<br>Uncomfortable<br>Neutral<br>Comfortable | 0 (0%)<br>5 (12.5%)<br>35 (87.5%) |
| Robotic system is acceptable to receive care<br>Disagree<br>Neutral<br>Agree | 1 (2.5%)<br>5 (12.5%)<br>34 (85%) |
| Willing to interact with the system again<br>Disagree<br>Neutral<br>Agree | 1 (2.5%)<br>2 (5%)<br>37 (92.5%) |

### Decision analytic model

Our decision analytic model found that the robotic system could prevent up to 2.68 infections (sensitivity analysis: 1.71 - 2.78 infections) over approximately one year per ED (Supplementary Table 3). Expanded across the United States as a method to facilitate contactless evaluations in the emergency department setting, we estimate the prevention of 16,000 infections per year if the system could prevent all cases of healthcare-acquired COVID.^24^ Additionally, the robotic strategy could be less expensive at $500-100,000 per year compared to staffing and equipping additional HCW at approximately $1 million per year (Supplementary Table 2). Furthermore, considering that one symptomatic COVID infection costs on average $3000, the robotic strategy could be significantly cost-saving.^25^

## Discussion

Risk of SARS-CoV-2 acquisition and enhanced social distancing measures have changed the way in which in-person healthcare visits are conducted during the COVID-19 pandemic. While robotic systems have been adopted for hospital-based tasks and surgical procedures, little work has been done to explore the deployment of patient-facing systems and their potential cost savings in the hospital. This investigation demonstrates that there is interest among the general public in accepting the use of robotic systems for patient interactions in the hospital, and this interest is reflected within a real-world pilot deployment of a robot to facilitate teletriage and patient interviewing in the ED during the COVID-19 pandemic. Among HCWs, adopting robotic systems to evaluate patients with COVID-19 symptoms is useful because it may decrease acquisition of COVID-19 by HCWs thereby preserving a crucial workforce during surges of infectious cases. Additionally, by minimizing exposure to patients, HCW can conserve crucial PPE. Additionally, we modeled the deployment of a robotic system in the emergency department for teletriage during the COVID-19 pandemic and demonstrated the potential public health and economic impact of the system via prevention of healthcare-acquired SARS-CoV-2 infection and conservation of HCW and PPE. Taken together, this investigation suggests that a robotic system to facilitate contactless teletriage in the emergency department is feasible, acceptable and could have a large public health impact during the COVID-19 pandemic and potentially inform strategies for other infectious disease settings.

Our national survey results demonstrate that individuals find robotic systems useful for in hospital patient interaction. We sought to define potential situations in which robots may be deployed to manage patients with COVID-19 disease. This includes an initial ED-based evaluation using teletriage and telemedicine for patient to physician interactions, acquisition of contactless vital signs, basic SARS-CoV-2 testing by obtaining nasal and oral swabs, initial medical testing and resuscitation with placement of IV catheters and phlebotomy, and finally in the critically ill patient, potential assistance with tasks like proning. We anticipate with changes in hospital practice, robotic systems can be developed to assist with these tasks, especially during surges of patients with potential COVID-19 infection.

While robotic systems have been implemented in hospitals to restock and deliver supplies, their use in facilitating human interactions has been limited.^26,27^ Some pilot studies have demonstrated feasibility of deploying a robotic platform for telerounding on inpatient units.^28,29^ Here, we were able to train emergency HCW in the operation of a robotic system and integrate it into our existing telehealth platform to facilitate contactless triage in the ED. Unlike inpatient services, the ED poses a unique challenge in navigating robotic systems through chaotic environments and interacting with patients in varying locations.^30^ Despite these challenges, participants were able to engage with our robotic teletriage platform; this was considered by 82.5% of individuals to be equivalent to in-person evaluation. Individuals also reported that they would be willing to engage with this robotic system in future visits. By designing a robotic platform and triage system that is acceptable to patients, we suspect that we can continue to engage ED patients at a time when pre-COVID-19 norms of in-person visits are less likely to occur.

We also demonstrated that a robotic system in the ideal scenario could prevent up to about 2.78 infections per ED per year and be less costly than standard triage. As regional surges of COVID-19 disease emerge, decreasing supplies of PPE can be partially mitigated by allowing robotic systems to interact with low risk individuals seeking hospital-based care. These individuals can be seen efficiently with a scripted triage screening history. Additionally, instead of requiring clinicians to don and doff PPE between patients, a robotic system can rotate through care areas and rapidly assess patients with potential COVID-19 symptoms, thereby improving emergency department throughput. Finally, a robotic teletriage system can be placed on standby and activated in response to patients who may only need a brief COVID-19 screening, allowing a clinician who may be already evaluating other patients in the emergency department to rapidly pivot to conduct teletriage, fulfilling hospital Emergency Medicine Treatment and Labor Act requirements.^31^ These advantages together may result in significant cost savings at the hospital and national level.

This study had several limitations. Regarding our national survey, we utilized a complex approach of sample matching and weight adjustment that was previously validated. Nevertheless, internet-based, non-probablistic, opt-in panels can have significant biases including the need to have access to the internet, and membership in an opt-in panel which may limit the generalizability of survey results. Our survey was administered through a national sampling of individuals living in the United States. Depending on personal experiences with the pandemic, respondents’ attitudes towards robotic systems may vary. Additionally, the demographic enrolled in the survey was largely Caucasian and well-educated. Next, we conducted our study at a single, large, urban, academic emergency department. We also experienced robust information technology support from our collaborators and hospital administration. The experiences deploying a complex robotic system like this in other centers may vary depending on availability of these resources. Third, we utilized a highly agile, mobile robotic system to facilitate telemedicine in this investigation. The user response to other robotic systems may vary. Fourth, we decontaminated Spot using hospital alcohol wipes which may be time- and resource-intensive. Future iterations of a cleaning system may include an on-board automated cleaning system that can be remotely triggered after a patient encounter and an ultraviolet enclosure to permit sterilization during storage. Fifth, our decision analytic model had several simplifications: We assumed maintenance and integration costs of the robot would be minimal compared to acquisition and personnel costs. Our analysis may be variable depending on the true costs of a robotic system. Additionally, we assumed no difference between triage accuracy between the robotic and human strategies. Further studies are needed to understand clinical outcomes of robotic-enabled triage. Finally, rates of hospital-acquired COVID-19 are still poorly understood and highly variable. Our estimates are derived from a range of hospital-based studies, but true rates of transmission will change during peaks and surges of COVID-19 infection, as may costs associated with infection. An updated analysis is warranted as we obtain deeper understanding of these risks and costs.

Overall, we demonstrated that interaction with robotic systems to facilitate traditional in-person history taking in the emergency department is feasible and acceptable to patients. In the COVID-19 pandemic era with increased emphasis on minimizing in-person contact and conserving PPE, adopting robotic systems to conduct in-person tasks may be acceptable to patients. There are several considerations surrounding the operation of these systems in the hospital. For example, our national survey suggests that individuals may find robots useful in facilitating key hospital tasks that have been traditionally done in person. This may inform the development of additional robotic systems that may help minimize exposure of HCW to individuals with COVID-19 disease while performing tasks like nasal swabs, IV placement, and assisting patients in bed. Because of the pilot nature of our investigation, we had on-site physician operators to control the robot. Future iterations of robotic telemedicine platforms may include remote operators like individuals who are considered at higher risk of complications from COVID-19 or those recovering from COVID-19. These additional sources of HCW may be instrumental in conducting assessments of lower risk individuals while able to work remotely as they recover or minimize their own exposure to SARS-CoV-2. Additionally, human controlled robotic systems still require training in maintenance and operation of the robot in addition to tasks associated with patient care. Developing fully autonomous robotic systems which can navigate around obstacles in a rapidly changing hospital environment may allow HCW who utilize these systems to focus on clinical patient care.

## Data Availability

Solidworks plans for healthcare payloads used on the quadruped robotic system (Boston Dynamics) in this investigation are available at https://github.com/boston-dynamics/bosdyn-hospital-bot. Individual data for the public survey, clinical study and decision analysis model is included as supplemental data to this manuscript.

## Declaration of interest

PRC is supported by NIH K23DA044874, R44DA051106, Hans and Mavis Lopater Psychosocial Foundation and e-ink corporation; JNC is supported by NIH T32DK007191-45; EWB is supported by NIH R01DA047236; GT is supported by the Karl van Tassel (1925) Career Development Professorship at MIT, the Department of Mechanical Engineering, MIT and Division of Gastroenterology, Brigham and Women’s Hospital. Complete details of all relationships for profit and not for profit for G.T. can be found at the following link: https://www.dropbox.com/sh/szi7vnr4a2ajb56/AABs5N5i0q9AfT1IqIJAE-T5a?dl=0. MD and MR are employees of Boston Dynamics

## Acknowledgements

The authors acknowledge the dedicated work of Andrew Tsang, Seth Davis, Gene Merewether, Joy Hui, Mike Grygorcewicz, Kim Ang and Nick Sipes from Boston Dynamics in providing technical support for this investigation.

## Supplementary

**Supplementary Figure 1.**
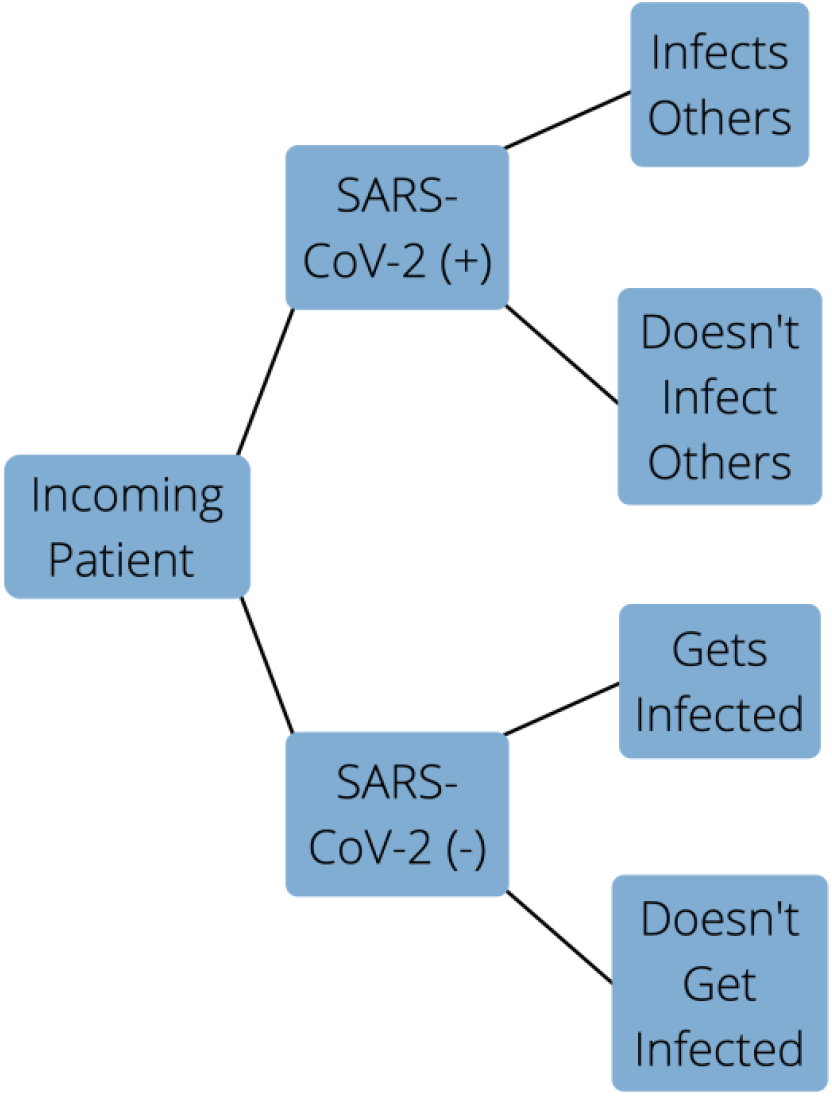
Schematic of the decision analytic model comparing robotic triage vs. standard (human) triage.

**Supplementary Table 2.**
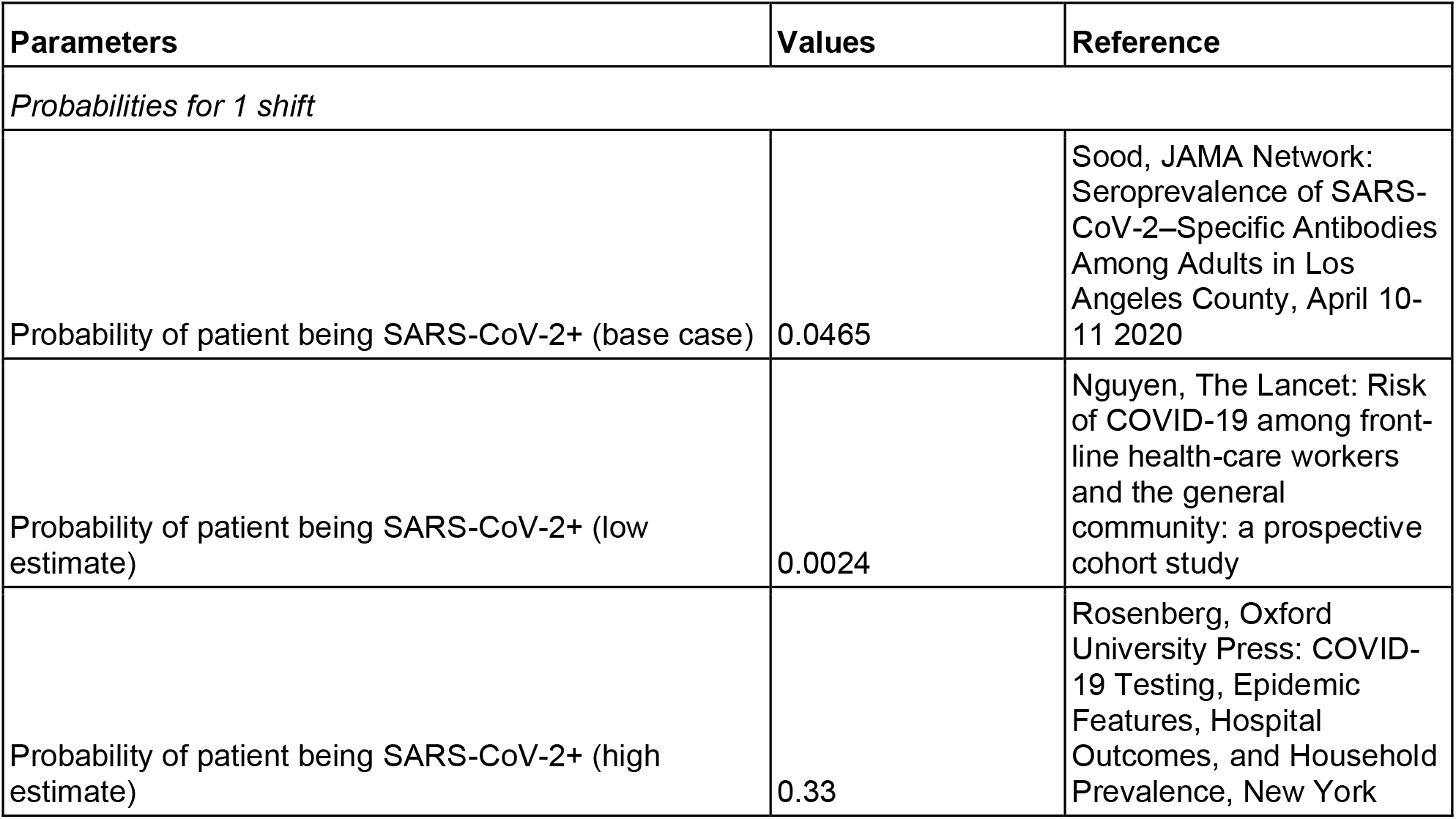
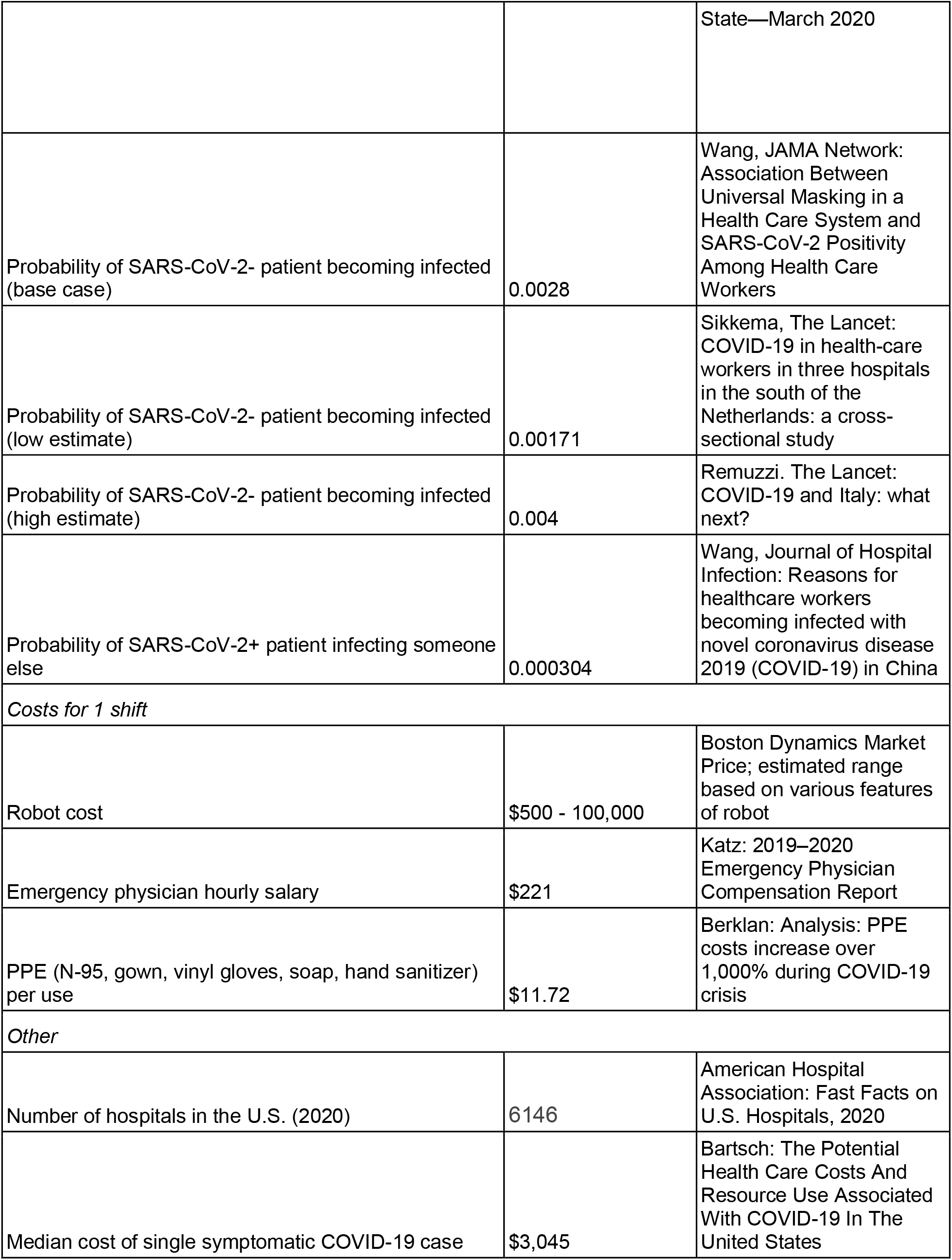
Parameters used in the decision tree.

**Supplementary Table 3.** Results from the decision analytic model.

|  | Infections avoided per year<br>per ED |
| --- | --- |
| Base case | 2.68 |
| Low estimate | 1.71 |
| High estimate | 2.78 |

## Notes

### Clinical Trial

NCT04452695

### Funding Statement

PRC supported by NIH K23DA044874, R44DA051106, Hans and Mavis Lopater Psychosocial Foundation and eink corporation. JNC is supported by NIH T32DK007191. EWB supported by NIH R01DA047236. GT is supported by the Karl van Tassel Career Development Professorship at MIT, the Department of Mechanical Engineering, MIT and Division of Gastroenterology, Brigham and Women's Hospital

### Author Declarations

Mass General Brigham Institutional Review (IRB) approval was received prior to initiating the study (MGB 2020P000957).

